# Tryptophan catabolites, inflammation, and insulin resistance as determinants of chronic fatigue syndrome and affective symptoms in Long COVID

**DOI:** 10.1101/2023.03.11.23287152

**Authors:** Hussein Kadhem Al-Hakeim, Anwar Khairi Abed, Shatha Rouf Moustafa, Abbas F Almulla, Michael Maes

## Abstract

**Background:** Critical COVID-19 disease is accompanied by depletion of plasma tryptophan (TRY) and increases in indoleamine-dioxygenase (IDO)-stimulated production of neuroactive tryptophan catabolites (TRYCATs), including kynurenine (KYN) and quinolinic acid. The TRYCAT pathway has not been studied extensively in association with the physiosomatic and affective symptoms of Long COVID.

**Methods:** In the present study, we measured serum tryptophan (TRY), TRYCATs, insulin resistance (using the HOMA2-IR index), C-reactive protein (CRP), physiosomatic, depression and anxiety symptoms in 90 Long COVID patients, 3-10 months after remission of acute infection.

**Results:** We were able to construct an endophenotypic class of severe Long COVID (22% of the patients) with very low TRY and oxygen saturation (SpO2, during acute infection), increased kynurenine, KYN/TRY ratio, CRP, and very high ratings on all symptom domains. One factor could be extracted from physiosomatic symptoms (including chronic fatigue-fibromyalgia), depression, and anxiety symptoms, indicating that all domains are manifestations of the common physio-affective phenome. Three Long COVID biomarkers (CRP, KYN/TRY, IR) explained around 40% of the variance in the physio-affective phenome. The latter and the KYN/TRY ratio were significantly predicted by peak body temperature (PBT) and lowered SpO2 during acute infection. One validated latent vector could be extracted from the three symptom domains and a composite based on CRP, KYN/TRY, IR (Long COVID), and PBT and SpO2 (acute COVID-19).

**Conclusion:** The physio-affective phenome of Long COVID is a manifestation of inflammatory responses during acute and Long COVID and lowered plasma tryptophan and increased kynurenine may contribute to these effects.

## Introduction

After the COVID-19 pandemic, which caused millions of acute cases and millions of deaths, healthcare professionals are facing another crisis due to the development and/or persistence of symptoms following the acute phase of SARS-CoV-2 infection (typically after three months), a condition known as Long COVID (Fernández-de-Las-Peñas 2022). Long COVID is most often defined as symptoms lasting more than three months after the onset of acute COVID-19 (Shah, Hillman et al. 2021, Yong 2021). The most frequently reported symptoms associated with long-term COVID are chronic fatigue, dyspnea, affective symptoms (anxiety and depression) and cognitive impairments (Arnold, Hamilton et al. 2021, Bliddal, Banasik et al. 2021, Davis, Assaf et al. 2021, Huang, Huang et al. 2021, Shah, Hillman et al. 2021, Van den Borst, Peters et al. 2021, Yong 2021, Guo, Benito Ballesteros et al. 2022). Within six months following the onset of the first COVID-19 symptom, about one-third of COVID-19 survivors experience neuropsychiatric symptoms, such as sleeplessness, anxiety, or depression (Taquet, Geddes et al. 2021).

Several pathways are believed to underlie the physio-affective phenome of Long COVID, which includes physiosomatic and affective (depression and anxiety) symptoms (Al-Hakeim, Al-Rubaye et al. 2022). Initially, the physio-affective phenome of chronic COVID was substantially predicted by reduced oxygen saturation (SpO2) and elevated peak body temperature (PBT) during the acute phase of SARS-CoV-2 infection (Al-Hadrawi, Al-Rubaye et al. 2022). This suggests that the Long COVID physio-affective phenome is the outcome of immune-inflammatory processes during the acute infection, at least in part (Al-Hadrawi, Al-Rubaye et al. 2022, Maes, Al-Rubaye et al. 2022, Al-Hakeim, Al-Rubaye et al. 2023, Al-Hakeim, Al-Rubaye et al. 2023). Second, the physio-affective phenome of Long COVID is associated with neuro-oxidative stress toxicity (OSTOX) and decreased antioxidant defenses (ANTIOX) (Maes 2011, Maes, Rachayon et al. 2022, Al-Hakeim, Al-Rubaye et al. 2023), activation of the NLRP3 inflammasome (Al-Hakeim, Al-Rubaye et al. 2022), inflammation as indicated by increased levels of C-reactive protein (CRP), and increased insulin resistance as assessed with HOMA2-IR index (Al-Hakeim, Al-Rubaye et al. 2023).

According to a recent meta-analysis, the acute infectious phase is characterized by a considerable activation of indoleamine-2,3-dioxygenase (IDO), which leads to tryptophan depletion and an increase in the synthesis of tryptophan catabolites (TRYCATs) (Almulla, Thipakorn et al. 2022). Moreover, activation of the TRYCAT pathway is related to critical illness and higher mortality (Almulla and Maes 2022). IDO is triggered by reactive oxygen species and pro-inflammatory cytokines, which are elevated during the acute infection phase (Maes, Galecki et al. 2011). These cytokines include interferon (IFN)-α, IFN-γ, interleukin (IL)-1β, tumor necrosis factor (TNF)-α, and IL-18 (Liebau, Baltzer et al. 2002, Oxenkrug 2007, Maes, Leonard et al. 2011). Nevertheless, tryptophan catabolites (TRYCATs) in long-COVID have not yet been investigated. Tryptophan depletion and increased synthesis of neuroactive TRYCATs may result from IDO activation. Several neurotoxic metabolites, including kynurenine (KYN), 3-hydroxykynurenine (3HK), anthranilic acid (AA), 3-hydroxyanthranilic acid (3-HA), and quinolinic acid (QA) are formed, but also TRYCATs with more protective properties are formed, such as kynurenic acid (KA) (Maes, Leonard et al. 2011, Almulla and Maes 2022). TRYCATs such as 3HK and QA, for instance, generate neuro-oxidative toxicity, resulting in oxidative cell damage and lipid peroxidation (Goldstein, Leopold et al. 2000, Smith, Smith et al. 2009, Reyes Ocampo, Lugo Huitrón et al. 2014, Almulla and Maes 2022). Tryptophan deficiency is also characteristic of major depression, somatization disorder, schizophrenia, cognitive deficits, and physiosomatic symptoms (Maes, Minner et al. 1991, Maes and Rief 2012, Almulla, Thipakorn et al. 2022, Almulla, Vasupanrajit et al. 2022). It is thought that increased production of neurotoxic TRYCATs may contribute to the neuropsychiatric symptoms of major depression, bipolar disorder, schizophrenia, anxiety, somatization disorder, and chronic fatigue syndrome, as well as neurocognitive impairments (Maes, Galecki et al. 2011, Cordeiro, Guimaraes et al. 2014, Morris, F Carvalho et al. 2016, Ormstad, Simonsen et al. 2020, Milaneschi, Allers et al. 2021). Nevertheless, no research has investigated whether the physio-affective phenome of Long COVID is associated with IDO activation above and beyond the effects of inflammation and IR.

Consequently, the objectives of the current study are to: a) examine the association between the long COVID physio-affective phenome and TRYCAT pathway activity; b) determine whether the TRYCAT pathway during long COVID is associated with inflammation during the acute infectious phase; and c) determine whether TRYCATs have an effect beyond that of CRP and IR.

## Subjects and Methods

### Subjects

We employed both a case-control research design (to explore differences between COVID-19 participants who developed mild versus severe Long COVID and those without Long COVID) and a retrospective cohort study design (to examine the effects of acute-phase biomarkers on Long COVID symptoms). In the last three months of 2022, we recruited 90 subjects 3-12 months after having suffered from acute COVID-19 infection, namely 60 Long COVID patients and 30 control subjects who did not develop Long COVID. The Long COVID patients were diagnosed using the officially published WHO criteria of post-COVID (long COVID) (WHO 2021), namely: a) they suffer from at least two symptoms that impair daily functioning, including fatigue, memory or concentration problems, shortness of breath, chest pain, persistent cough, difficulty speaking, muscle aches, loss of smell or taste, affective symptoms, cognitive impairment, or fever; b) they have a history of proven SARS-CoV-2 infection, and c) the symptoms persisted beyond the acute phase of illness or are manifested during recovery from acute COVID-19 infection, lasted at least two months and are present 3-4 months after the onset of COVID-19 (WHO 2021). The 30 controls did not comply with any of these criteria, except that they had a proven history of acute SARS-CoV-2 infection. Thus, the controls exhibited no clinical indications of acute infection, such as dry cough, sore throat, shortness of breath, lack of appetite, flu-like symptoms, fever, night sweats, or chills. During the acute phase of the illness, all participants were admitted to one of the official quarantine facilities in Al-Muthanna General Hospital and Al-Khidr General Hospital, at Al-Muthanna Governorate, Iraq. Senior doctors and virologists made the diagnosis of SARS-CoV-2 infection and acute COVID-19 based on a) presence of an acute respiratory syndrome and the disease’s standard symptoms of fever, breathing difficulties, coughing, and loss of smell and taste; b) positive reverse transcription real-time polymerase chain reaction findings (rRT-PCR); and c) positive IgM directed to SARS-CoV-2. We included only participants who tested negative for rRT-PCR 3-12 months afer the acute infection. We selected the controls so that about one-third of them had distress or adjustment symptoms as a result of lockdowns and social isolation to account for their confounding effects, which are also seen in Long COVID patients. Thus, one-third of the controls showed Hamilton Depression Rating Scale (HAMD) (Hamilton 1960) values between 7 and 10. COVID patients and controls were excluded if they had a lifetime history of psychiatric axis-1 disorders, including major affective disorders such as major depression and bipolar disorder, dysthymia, generalized anxiety disorder and panic disorder, schizo-affective disorder, schizophrenia, psycho-organic syndrome, and substance use disorders except tobacco use disorder (TUD). Moreover, we excluded patients and controls who suffered from neurodegenerative and neuroinflammatory disorders, such as chronic fatigue syndrome, Parkinson’s or Alzheimer’s disease, multiple sclerosis, stroke, or systemic (auto)immune diseases such as diabetes mellitus, psoriasis, rheumatoid arthritis, inflammatory bowel disease and scleroderma, and liver and renal disease. Additionally, pregnant and breastfeeding women were omitted from this study.

All controls and patients, or their parents/legal guardians, gave written informed permission before participation in the research. The research was approved by the University of Kufa’s institutional ethics board (8298/2022). The study was conducted under Iraqi and international ethical and privacy laws, including the World Medical Association’s Declaration of Helsinki, The Belmont Report, the CIOMS Guideline, and the International Conference on Harmonization of Good Clinical Practice; our institutional review board adheres to the International Guideline for Human Research Safety (ICH-GCP).

### Clinical measurements

A senior psychiatrist used a semi-structured interview to collect socio-demographic and clinical data from controls and Long COVID patients three to four months after the acute infectious phase of COVID-19. Three to four months after the onset of acute COVID-19, a senior psychiatrist assessed the following rating scales: a) the 12-item Fibro-Fatigue (FF) scale to assess chronic fatigue and fibromyalgia symptoms (Zachrisson, Regland et al. 2002); b) the HAMD to assess the severity of depression (Hamilton 1960); and c) the Hamilton Anxiety Rating Scale (HAMA) (Hamilton 1959) to assess the severity of anxiety. We calculated: a) key HAMD depressive symptoms (pure HAMD) as the sum of sad mood + feelings of guilt + suicidal thoughts + loss of interest; b) key HAMA anxiety symptoms (pure HAMA), which were defined as anxious mood + tension + fears + anxiety behavior during the interview; and c) physiosomatic symptoms (physiosom symptoms) as a z composte score based on HAMD items, such as somatic anxiety + gastrointestinal (GIS) anxiety + genitourinary anxiety + hypochondriasis, HAMA items, such as somatic sensory + cardiovascular + GIS + genitourinary + autonomic symptoms (respiratory symptoms were not included in the sum) and FF symptoms, such as muscular pain + muscle tension + fatigue + autonomous symptoms + gastrointestinal symptoms + headache + a flu-like malaise (thus excluding all cognitive and affective symptoms). It is of paramount importantance to consider both physiosomatic and affective symptom domains because the rating scales used here contain both symptom domains. Not considering both symptom domains would not allow us to draw any conclusions on depression and anxiety in relation to the TRYCAT pathway. To construct a score reflecting the severity of the physio-affective phenome, we extracted the first factor from the pure FF and pure and physiosom HAMA and HAMD scores (labeled: the physio-affective phenome) (Al-Hadrawi, Al-Rubaye et al. 2022, Al-Jassas, Al-Hakeim et al. 2022). Additionally, we constructed z unit-based composite scores indicating all Fibro-Fatigue symptoms (muscle pain + cramps + fatigue), sleep problems, and cognitive symptoms using all relevant HAMD, HAMA, and FF items (z transformed).

We recorded the immunizations received by each participant, namely AstraZeneca, Pfizer, or Sinopharm. TUD was diagnosed using DSM-5 criteria. We determined the body mass index (BMI) by dividing the body weight in kilograms by the square of the height in meters.

### Assays

Fasting blood samples were taken in the early morning between 7.30-9.00 a.m. after awakening and before having breakfast. Five milliliters of venous blood were drawn and transferred into clean plain tubes. Hemolyzed samples were rejected. After ten minutes, the clotted blood samples were centrifuged for five minutes at 3,000 rpm, and then serum was separated and transported into three new Eppendorf tubes until assay. Serum tryptophan, kynurenine, and insulin were measured using ELISA kits supplied by Nanjing Pars Biochem Co., Ltd. (Nanjing, China). Serum 3HK was estimated by ELISA kits supplied by Melsin Medical Co, (Jilin, China). ELISA kits for the estimation of KA and QA were supplied by ELK Biotech. Co., Ltd (Wuhan, China). All kits have an inter-assay CV of less than 10%. CRP latex slide tests (Spinreact^®^, Barcelona, Spain) were utilized for CRP measurements in human serum. Serum fasting blood glucose (FBG) was measured spectrophotometrically using a ready-for-use kit supplied by Biolabo^®^ (Maizy, France).

A well-trained paramedical specialist measured SpO2 using an electronic oximeter supplied by Shenzhen Jumper Medical Equipment Co. Ltd., and a digital thermometer was used to assess peak body temperature (PBT) (sublingual). We collected these indicators from patient records and analyzed the lowest SpO2 and peak body temperature values recorded during the acute phase of the illness. We created a new indicator based on these two evaluations that represents the severity of inflammation during the acute phase as z PBT – z SpO2. Consequently, we computed some composite scores: a) the acute phase of infection: z PBT + z duration of acute infection – z SpO2), b) insulin resistance: z FBG + z insulin (z IR), c) IDO activity: z KYN – zTryptophan (z KYN/TRY) and the sum of all z transformed TRYCATs, d) a composite based on z KYN/TRY + z CRP + z IR (LCB, Long COVID biomarkers); and e) a composite based on acute and Long COVID biomarkers (ALCB): z KYN/TRY + z CRP + z IR + z PBT – z SpO2.

### Statistics

Analysis of variance (ANOVA) was performed to determine if there were variations in scale variables across groups, and analysis of contingency tables was employed to determine connections between nominal variables. We used Pearson’s product-moment correlation coefficients to examine relationships between biomarkers and SpO2, PBT, and clinical physio-affective assessments. We employed a univariate general linear model (GLM) analysis to characterize the associations between classifications and biomarkers while accounting for confounding factors such as TUD, sex, age, BMI, and education. Additionally, we obtained model-generated estimated marginal means (SE) values and used protected pairwise comparisons between group means using Fisher’s least significant difference. Multiple comparisons were examined using a p-correction for false discovery rate (FDR) (Benjamini and Hochberg 1995). Multiple regression analysis was used to determine the important TRYCATs, other biomarkers, or cofounders that predict physio-affective measures. We used an automated stepwise technique with an 0.05 p-value to enter and p=0.06 to remove. We calculated the standardized beta coefficients for each significant explanatory variable using t statistics with exact p-value, as well as the model F statistics and total variance explained (R^2^) as effect size. Additionally, we examined multicollinearity using the variance inflation factor VIF and tolerance. We checked for heteroskedasticity using the White and modified Breusch-Pagan tests, and where necessary, we generated parameter estimates with robust errors. The tests were two-tailed, and statistical significance was defined as a p-value of 0.05. Based on clinical and biomarker analyses, a two-step cluster analysis was done to generate patient clusters. Using a silhouette measure of cohesiveness and separation of more than 0.5, the cluster solution was accepted. Using principal component analysis (PCA), we extracted a first PC that could be validated as a construct if the variance explained is greater than 50.0%, all loadings on the first PC are greater than 0.70, and the anti-image correlation matrix, Kaiser-Meyer-Olkin (KMO), and Bartlett’s sphericity test are satisfactory. We used IBM SPSS 28 for Windows to examine the data. With an effect size of 0.2, alpha of 0.05, power of 0.8, and five predictors, an a priori power analysis (G*Power 3.1.9.4) determined that the minimum sample size for multiple regression analysis should be 70.

## Results

### Results of PCA and cluster analysis

We were able to extract one PC from the pure depression and anxiety scores and physiosomatic scores as well (KMO=0.820, Bartlett’s test of sphericity: χ2=136.85, df=3, p<0.001, variance explained = 75.55%, all loadings on the first PC are > 0.810). As such, this first PC reflects the severity of the physio-affective phenome of Long COVID. The participants were divided into those without any symptoms (No Long COVID) and patients with Long-COVID divided into two groups: severe Long COVID and less severe groups (Long COVID). Moreover, we were able to extract one PC from the pure depression and anxiety and physiosomatic scores and the ALCB (KMO=0.702, Bartlett’s test of sphericity: χ2=180.77, df=6, p<0.001, variance explained = 69.11% and all loadings on the first PC > 0.770). Using two-step cluster analysis performed on the pure depression, anxiety and physiosomatic scores, CRP, z IR, z KYN/TRY, SpO2, PBT, and the diagnosis controls versus patients, as a categorial variable, we constructed three clusters (the silhouette measure of cohesion and separation was 0.61 indicating an adequate cluster solution). **Table 1** shows the features of these three clusters.

**Table 1.**
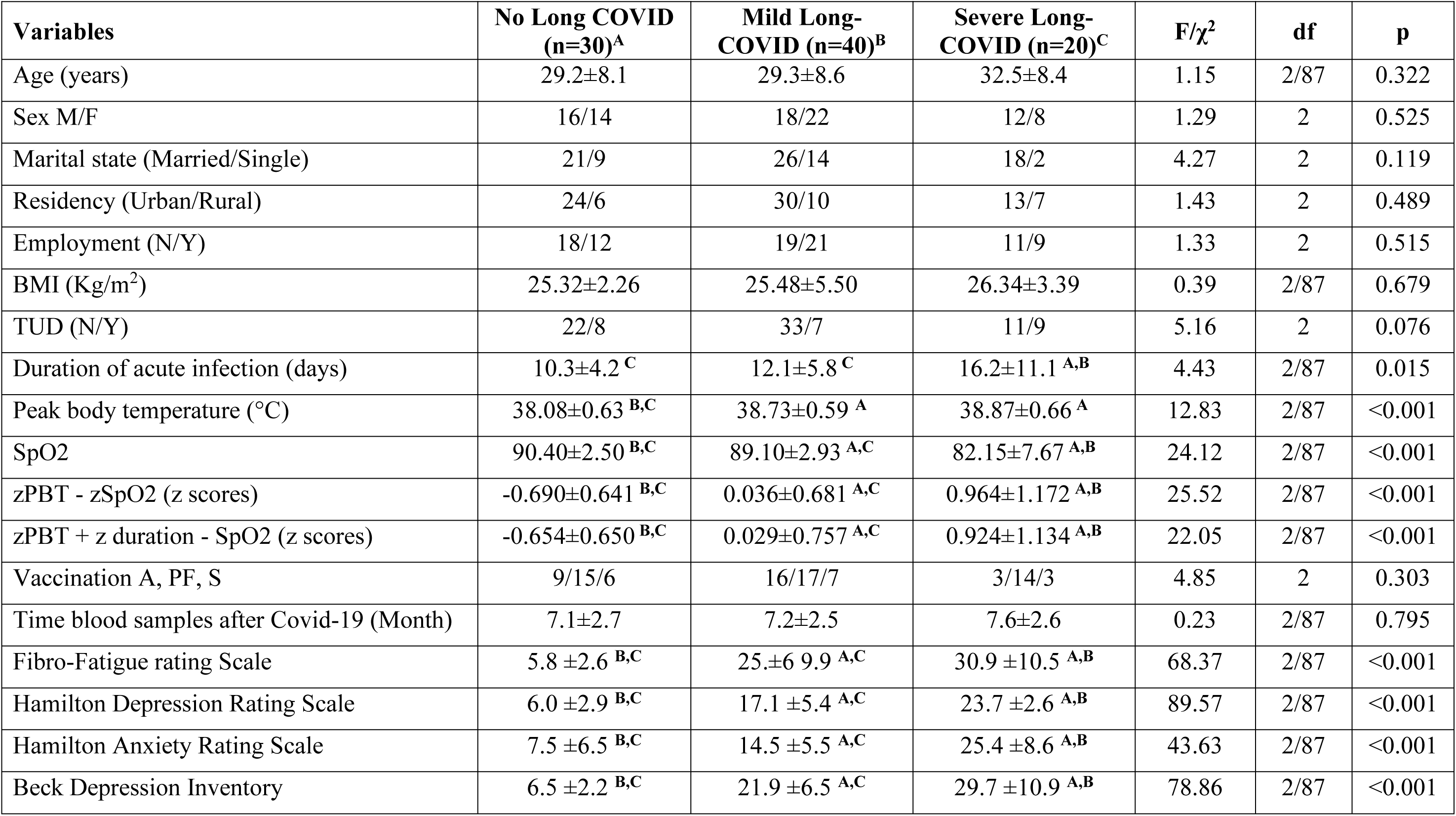
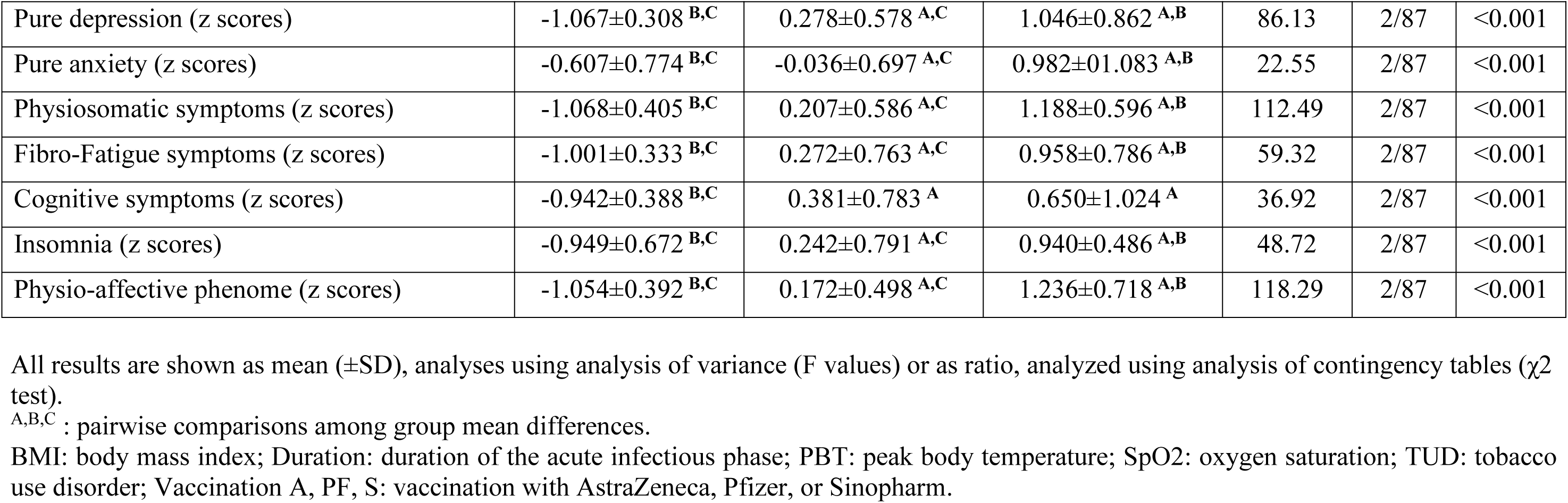
Demographic and clinical data in subjects, 3-12 months after suffering from acute COVID-19, divided into those without any symptoms of Long COVID (No Long COVID), and Long-COVID patients divided into two clusters, namely those with severe (severe Long COVID) and less severe (mild Long COVID) illness.

### Sociodemographic and clinical data

Table 1 shows that there were no significant differences among groups in age, sex ratio, marital status, residency, employment, BMI, TUD, vaccination type, and time of blood sampling after COVID-19. The duration of the acute infection phase was greater in severe Long-COVID than in mild Long-COVID and control groups with the lowest duration in the last group. The severe Long-COVID group showed a significantly higher PBT than mild Long-COVID and controls. During the acute phase of COVID infection, SpO2 was lower in the severe Long COVID group compared with the mild Long COVID followed by the control group. Pure depression, anxiety, and physiosomatic symptoms, fibro-fatigue symptoms, insomnia, and the physio-affective phenome scores, SpO2, z PBT – z SpO2, and z PBT + z duration – z SpO2 were significantly different between the three groups, and increased from controls → mild Long COVID → severe Long COVID. PBT and cognitive symptoms were higher in patients than in controls.

### Biomarkers

The biomarkers assessments in the three study groups are presented in **Table 2**. There was no significant difference among the three study groups in serum KYN, KA, QA, 3-HK, and the composite of all 4 TRYCATs. Severe Long COVID showed significantly higher CRP, LCB, and ALCB scores as compared with the other groups with the lowest value in controls. FBG was significantly higher in the severe Long COVID group compared with controls. Tryptophan is significantly lower in the severe Long COVID group compared with the mild Long COVID group. Serum insulin is higher in patients than in controls. The z KYN/TRY composite is higher in severe Long COVID than in both other groups.

**TABLE 2.** Biomarker measurements in subjects 3-12 months after suffering from acute COVID-19, divided into those without any symptoms of Long COVID (No Long COVID), and Long-COVID patients divided into two clusters, namely those with severe (severe Long COVID) and less severe (mild Long COVID) illness.

| Variables | No Long COVID<br>(n=30)<sup>A</sup> | Mild Long-COVID<br>(n=40)<sup>B</sup> | Severe Long-COVID (n=20)<sup>C</sup> | F (df=2/87) | p |
| --- | --- | --- | --- | --- | --- |
| C-Reactive Protein (CRP) (mg/l) | 6.22±2.51 <sup>B, C</sup> | 11.39±7.08 <sup>A, C</sup> | 16.53±9.06 <sup>A, B</sup> | 18.81 | <0.001 |
| Fasting Blood Glucose (FBG) (mM) | 5.73±0.52 <sup>C</sup> | 5.97±0.59 | 6.27±0.56 <sup>A</sup> | 5.49 | 0.006 |
| Insulin (pM) | 64.69±15.05 <sup>B, C</sup> | 76.97±25.95 <sup>A</sup> | 82.55±22.88 <sup>A</sup> | 4.49 | 0.014 |
| z FBG + z Insulin (z IR) (z score) | -0.530±0.762 <sup>B, C</sup> | 0.102±0.948 <sup>A</sup> | 0.591±1.058 <sup>A</sup> | 9.40 | <0.001 |
| L-Tryptophan (TRY) (ng/ml) | 11.65±3.96 | 12.93±6.47 <sup>C</sup> | 8.81±2.28 <sup>B</sup> | 5.32 | 0.007 |
| 3-OH-Kynurenine (ng/mL) | 2.22±2.13 | 2.64±2.03 | 2.72±1.76 | 0.51 | 0.605 |
| Quinolinic acid (ng/mL) | 7.06±3.84 | 6.46±3.60 | 5.70±3.97 | 0.79 | 0.456 |
| Kynurenic acid (ng/mL) | 266.2±101.1 | 247.2±93.6 | 237.9±105.5 | 0.56 | 0.574 |
| Kynurenine (KYN) (ng/mL) | 3.371±2.089 | 3.120±1.591 <sup>C</sup> | 4.426±2.774 <sup>B</sup> | 2.75 | 0.070 |
| z KYN – z TRY (z KYN/TRY) (z score) | -0.104±0.889 <sup>C</sup> | -0.302±0.876 <sup>C</sup> | 0.760±1.043 <sup>A, B</sup> | 9.20 | <0.001 |
| Composite all 4 TRYCATs (z score) | 0.040±1.083 | -0.072±0.899 | 0.083±1.102 | 0.19 | 0.826 |
| z CRP + z KYN/TRY + z IR (z score)) | -0.693±0.591 <sup>B, C</sup> | -0.022±0.817 <sup>A, C</sup> | 1.083±0.887 <sup>A, B</sup> | 32.19 | <0.001 |
| z CRP + z KYN/TRY + z IR + z PBT – z SpO2 (z score) | -0.845±0.567 <sup>B, C</sup> | 0.005±0.675 <sup>A, C</sup> | 1.257±0.685 <sup>A, B</sup> | 64.07 | <0.001 |

All results are shown as mean (±SD), analyses using analysis of variance (F values)
<sup>A,B,C</sup> : pairwise comparisons among group mean differences; TRYCATs: tryptophan catabolites; PBT: peak body temperature; SpO2: oxygen saturation

### Intercorrelation matrix among physio-affective symptoms and biomarkers

The intercorrelation matrix of selected symptom domains (fibro-fatigue, pure anxiety, pure depression, and physiosomatic) and biomarkers are presented in **Table 3**. The four clinical scores were significantly correlated with indicants of acute COVID-19 infection, CRP, and IR. There were significant associations between tryptophan (inversely) and the z KYN/TRY composite (positively) and fibro-fatigue, pure anxiety and physiosomatic (but not pure depression) symptoms.

**Table 3.** Intercorrelation matrix between physio-affective symptom domains and biomarkers and acute COVID-19 and Long COVID.

| Variables | z PBT – z SpO2 | Fibro-Fatigue | Pure Anxiety | Pure depression | Physiosomatic |
| --- | --- | --- | --- | --- | --- |
| Fibro-Fatigue | 0.408(<0.001) | 1 | 0.604(<0.001) | 0.701(<0.001) | 0.911(<0.001) |
| Pure Anxiety | 0.241(0.022) | 0.604(<0.001) | 1 | 0.428(<0.001) | 0.701(<0.001) |
| Pure Depression | 0.345(<0.001) | 0.701(<0.001) | 0.428(<0.001) | 1 | 0.756(<0.001) |
| Physiosomatic symptoms | 0.445(<0.001) | 0.911(<0.001) | 0.701(<0.001) | 0.756(<0.001) | 1 |
| Peak Body Temperature (PBT) | - | 0.316(0.002) | 0.167 | 0.294(0.005) | 0.375(<0.001) |
| Oxygen saturation (SpO2) | - | -0.290(0.006) | -0.221(0.036) | -0.222(0.035) | -0.370(<0.001) |
| Z PBT – z SpO2 | - | 0.375(<0.001) | 0.240(0.001) | 0.320(0.002) | 0.461(<0.001) |
| Z PBT +z Duration + z SpO2 | - | 0.438(<0.001) | 0.325(0.002) | 0.276(0.008) | 0.514(<0.001) |
| C-reactive protein (CRP) | 0.176(0.096) | 0.359(<0.001) | 0.288(0.006) | 0.567(<0.001) | 0.404(<0.001) |
| Z glucose + z insulin (z IR) | 0.136(0.200) | 0.368(<0.001) | 0.232(0.028) | 0.356(<0.001) | 0.377(<0.001) |
| Tryptophan (TRY) | -0.207(0.051) | -0.277(0.008) | -0.305(0.003) | -0.153(0.151) | -0.238(0.024) |
| Kynurenine (KYN) | 0.199(0.060) | 0.048(0.652) | 0.222(0.035) | 0.051(0.635) | 0.138(0.194) |
| z KYN/TRY | 0.273(0.009) | 0.237(0.025) | 0.384(<0.001) | 0.148(0.163) | 0.274(0.009) |
| All 4 TRYCATs | 0.088(0.407) | -0.047(0.659) | 0.108(0.311) | -0.152(0.152) | -0.002(0.985) |
| z CRP + z KYN/TRY + z IR | 0.334(0.001) | 0.499(<0.001) | 0.468 0.001 | 0.554 0.001 | 0.546 0.001 |
| z CRP + z KYN/TRY + z IR + z PBT – z SpO2 | - | 0.541(<0.001) | 0.445(<0.001) | 0.547(<0.001) | 0.620(<0.001) |

IR: insulin resistance, KYNTRYCATs: tryptophan catabolites; Duration: duration of the acute infectious phase; PBT: peak body temperature; SpO<sub>2</sub>: oxygen saturation;

This table shows that the severity of the acute infectious phase (z PBT – SpO2) is significantly correlated with all 4 symptom domains, z KYN/TRY, and LCB. The latter composite and ALCB were strongly associated with the four symptom domains as shown in Table 3. Moreover, the duration of the acute infectious phase was significantly correlated with the fibro-fatigue (r=0.339, p=0.001, n=90), pure anxiety (r=0.281, p=0.007), and physiosomatic (r=0.390, p<0.001) symptoms and z PBT – z SpO2 (r=0.335, p=0.001).

### Results of the multiple regression analysis

**Table 4** presents the results of multiple regression analyses with physio-affective symptom domains as dependent variables and biomarkers as explanatory variables. Regression #1 shows that 25.0% of the variance in the fibro-fatigue symptom score was explained by the regression on z IR, CRP, and z KYN/TRY. **Figure 1** shows the partial regression of the fibro-fatigue score on the LCB composite. Regression #2 shows that 20.6% of the variance in the cognitive score was explained by the regression on CRP and z IR. Regression #3 shows that 23.9 % of the variance in insomnia score was explained by the regression on CRP, z KYN/TRY, and z IR. Regression #4 shows that a considerable part of the variance in pure anxiety (23.9%) was explained by the regression on z KYN/TRY and CRP. **Figure 2** shows the partial regression of the pure anxiety score on the z KYN/TRY ratio. Regression #5 shows that 41.2% of the variance in the pure depression score was explained by CRP, z IR, and z KYN/TRY (all positively associated) and sex (higher in females). A significant part of the variance of the physiosomatic symptoms score (33.6%) can be explained by the regression on CRP, z KYN/TRY, z IR, and age as seen in Regression #6. Regression #7 shows that 41.0 % of the variance in the physio-affective phenome was explained by the regression on age and LCB. **Figure 3** shows the partial regression of the physio-affective phenome in long-COVID patients on LCB.

**Figure 1.**
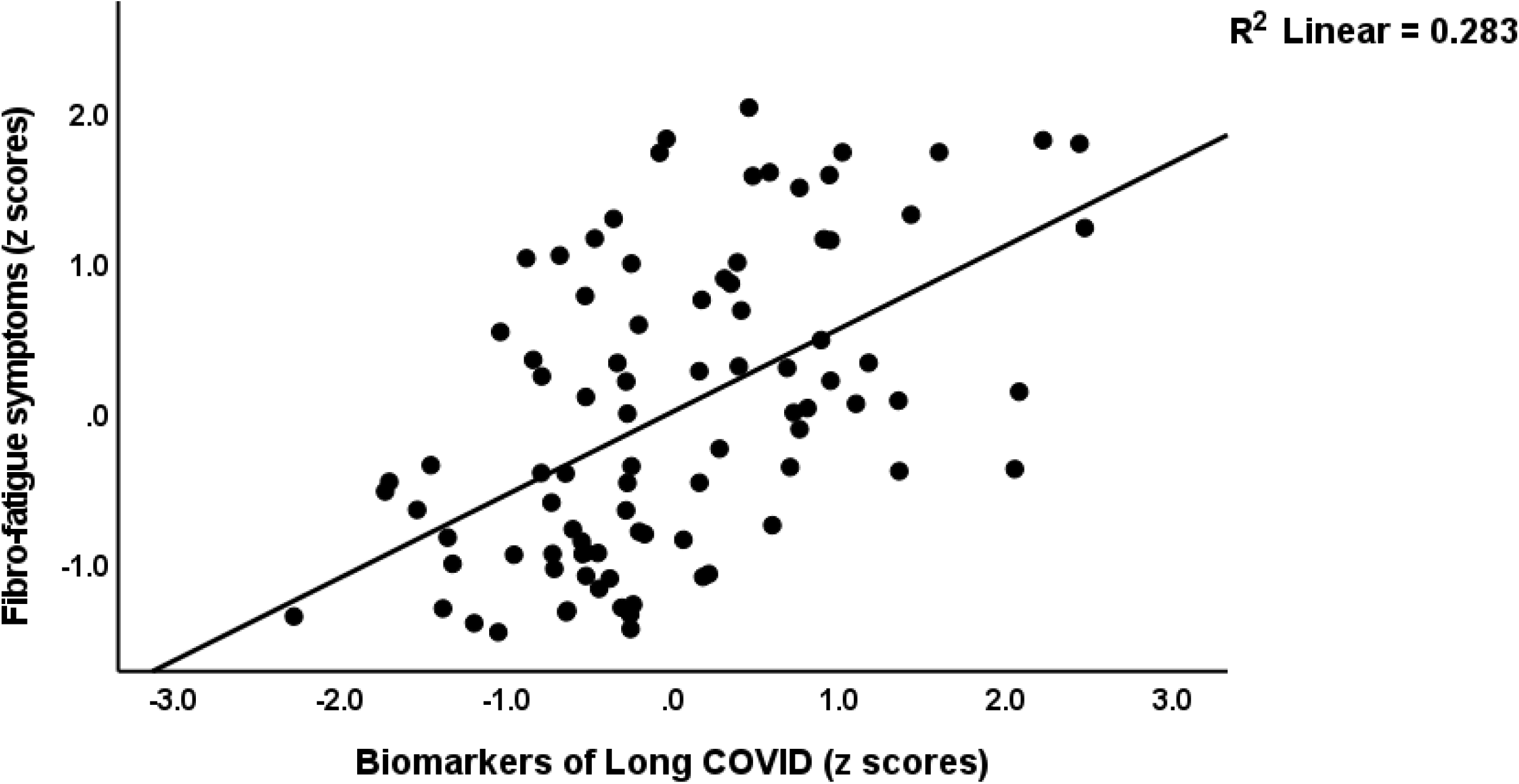
Partial regression of the fibro-fatigue score on the biomarkers of Long COVID (a composite based on C-reactive protein, the kynurenine/tryptophan ratio, and insulin resistance).

**Figure 2.**
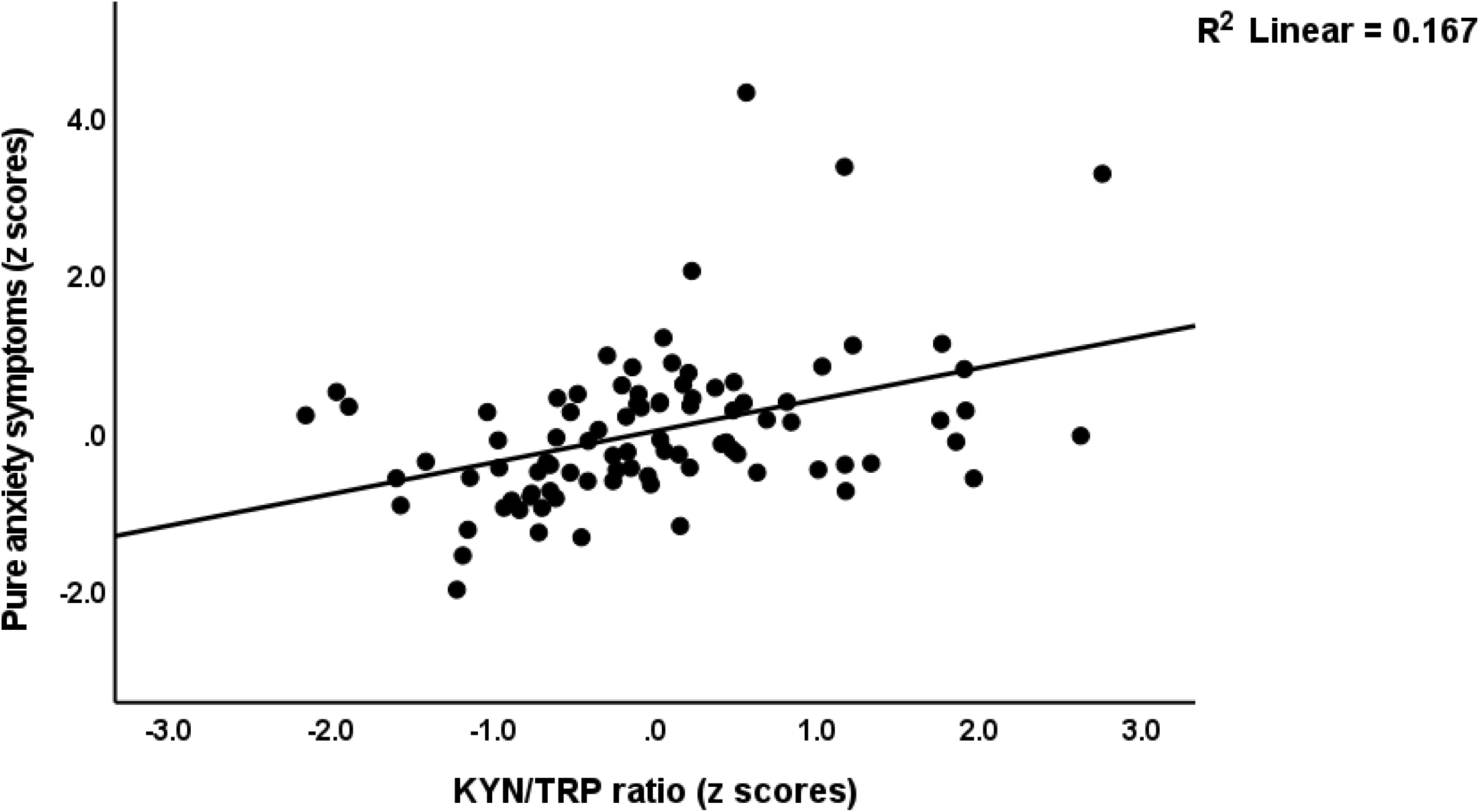
Partial regression of the pure anxiety score on kynurenine/tryptophan (KYN/TRY) ratio

**Figure 3.**
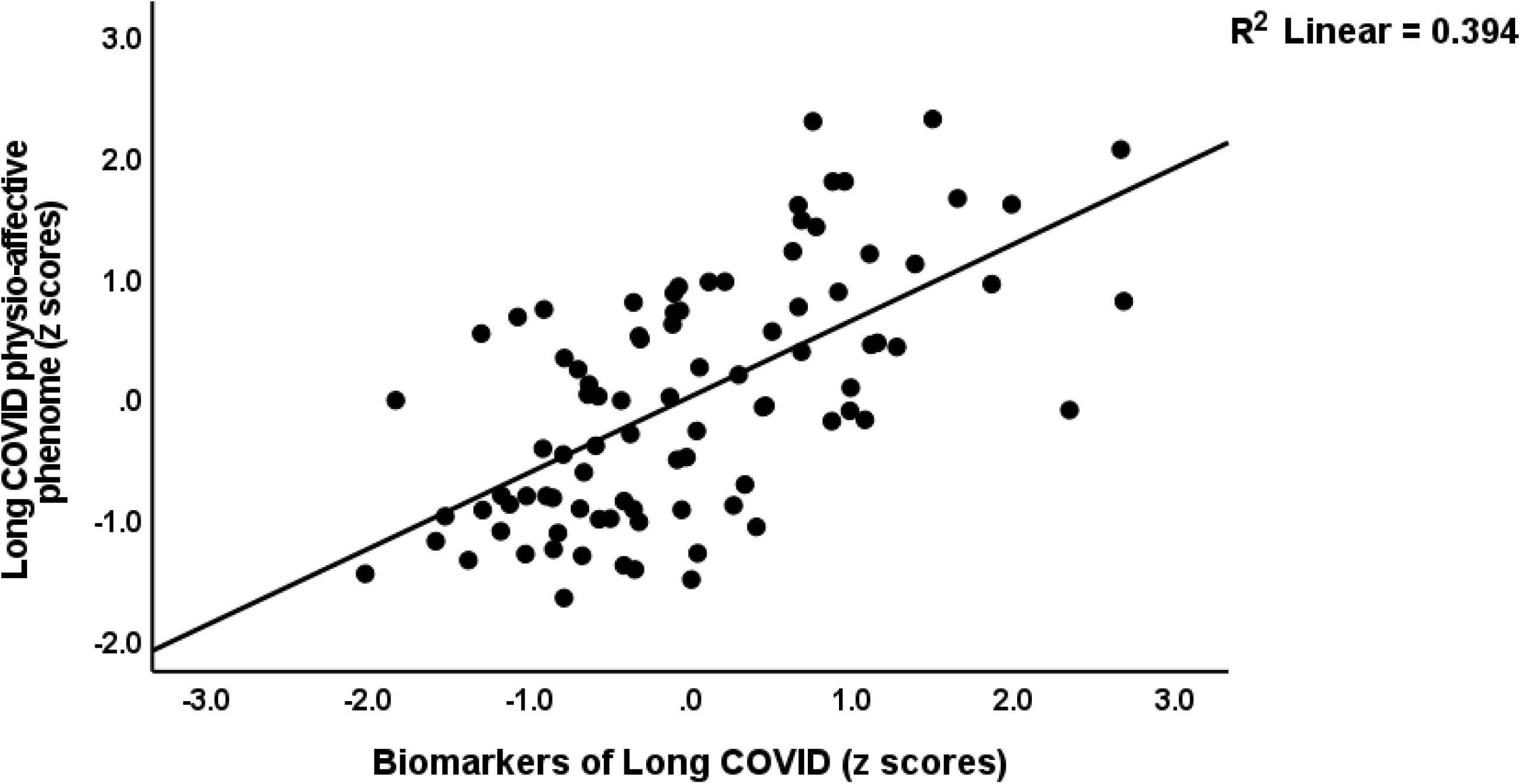
Partial regression of the physio-affective phenome of Long COVID on the biomarkers of that condition (a composite based on C-reactive protein, the kynurenine/tryptophan ratio, and insulin resistance).

**TABLE 4.** Results of multiple regression analysis with physio-affective symptom domains as dependent variables and biomarkers as explanatory variables

| Dependent Variables | Explanatory Variables | Coefficients of input variables |  |  | Model statistics |  |  |  |
| --- | --- | --- | --- | --- | --- | --- | --- | --- |
| | | $\beta$ | t | p | R<sup>2</sup> | F | df | p |
| #1. Fibro-Fatigue | Model |  |  |  | 0.250 | 9.57 | 3/86 | <0.001 |
|  | z IR | 0.259 | 2.605 | 0.011 |  |  |  |  |
|  | CRP | 0.280 | 2.811 | 0.006 |  |  |  |  |
|  | z KYN/TRY | 0.231 | 2.464 | 0.016 |  |  |  |  |
| #2. Cognitive deficits | Model |  |  |  | 0.206 | 11.25 | 2/87 | <0.001 |
|  | CRP | 0.283 | 2.781 | 0.007 |  |  |  |  |
|  | z IR | 0.271 | 2.668 | 0.009 |  |  |  |  |
| #3. Insomnia | Model |  |  |  | 0.239 | 9.00 | 3/86 | <0.001 |
|  | CRP | 0.331 | 3.304 | 0.001 |  |  |  |  |
|  | z KYN/TRY | 0.202 | 2.137 | 0.035 |  |  |  |  |
|  | z IR | 0.205 | 2.041 | 0.044 |  |  |  |  |
| #4. Pure Anxiety | Model |  |  |  | 0.239 | 13.66 | 2/87 | <0.001 |
|  | Z KYN/TRP | 0.395 | 4.224 | <0.001 |  |  |  |  |
|  | CRP | 0.302 | 3.231 | 0.002 |  |  |  |  |
| #5. Pure depression | Model |  |  |  | 0.412 | 14.88 | 4/85 | <0.001 |
|  | CRP | 0.533 | 5.9902 | <0.001 |  |  |  |  |
|  | z IR | 0.205 | 280 | 0.025 |  |  |  |  |
|  | Sex | -0.197 | -2.288 | 0.025 |  |  |  |  |
|  | z KYN/TRY | 0.179 | 2.133 | 0.036 |  |  |  |  |
| #6. Physiosomatic symptoms | Model |  |  |  | 0.336 | 10.74 | 4/85 | <0.001 |
|  | CRP | 0.315 | 3.338 | 0.001 |  |  |  |  |
|  | z KYN/TRY | 0.293 | 3.278 | 0.002 |  |  |  |  |
|  | z IR | 0.284 | 2.964 | 0.004 |  |  |  |  |
|  | Age | 0.191 | 2.112 | 0.038 |  |  |  |  |
| #7. Physio-affective phenome | Model |  |  |  | 0.410 | 29.09 | 2/87 | <0.001 |
|  | z CRP + z KYN/TRY + z IR | 0.632 | 7.528 | <0.001 |  |  |  |  |
|  | Age | 0.195 | 2.326 | 0.022 |  |  |  |  |

CRP: C-reactive protein, FBG: fasting blood glucose, IR: insulin resistance, SpO2: Saturation of Peripheral Oxygen, PBT: peak body temperature, KYN: kynurenine, TRY: tryptophan, sex: dummy variable with 0=female, 1=male

**Table 5** shows the results of the multiple regression analyses with physio-affective symptom domains as dependent variables and the biomarkers of Long COVID (z IR, z KYN/TRY, and CRP) and acute COVID-19 infection (SpO2, PBT, and duration of infection) as explanatory variables. Regression #1 revealed that 32.1% of the variance in the fibro-fatigue score can be explained by the regression on z PBT + z duration -z SpO2, CRP, and z IR. **Figure 4** shows the partial regression of the fibro-fatigue score on a composite based on all acute and Long COVID biomarkers. In regression #2, 27.0% of the variance in the cognition score was explained by the regression on CRP, PBT, and z IR. Regression #3 indicates that the regression on CRP and z PBT – z SpO2 can explain 23.9% of the variance of the insomnia score. According to regression #4, z KYN/TRY, CRP, z duration, and BMI may account for 36.5% of the variation in the pure anxiety score. Regression #5 found that zPBT – z SpO2 and CRP explain together 37.0% of the variation in the pure depression score. The regression on CRP, z PBT + z duration – z SpO2, and z KYN/TRY explained 41.1% of the variance in the physiosomatic symptoms score. Regression #7 shows that the regression on CRP, z PBT + z duration – z SpO2, and z KYN/TRY can explain 42.8% of the variation in the physio-affective phenome score. **Figure 5** shows that a large part of the variance in the physio-affective phenome is predicted by the cumulative effects of biomarkers of acute and Long COVID.

**TABLE 5.** Results of multiple regression analysis with physio-affective symptom domains as dependent variables and biomarkers, peak body temperature (PBT), and peripheral blood oxygen saturation (SpO2) as explanatory variables

| Dependent Variables | Explanatory Variables | Coefficients of input variables |  |  | Model statistics |  |  |  |
| --- | --- | --- | --- | --- | --- | --- | --- | --- |
| | | $\beta$ | t | p | R<sup>2</sup> | F | df | p |
| #1. Fibro-Fatigue | **Model** |  |  |  | 0.321 | 13.58 | 3/86 | <0.001 |
|  | z PBT + z duration -z SpO2 | 0.361 | 3.964 | <0.001 |  |  |  |  |
|  | CRP | 0.231 | 2.438 | 0.017 |  |  |  |  |
|  | Z IR | 0.219 | 2.285 | 0.025 |  |  |  |  |
| #2. Cognition | **Model** |  |  |  | 0.270 | 10.59 | 3/86 | <0.001 |
|  | CRP | 0.240 | 2.424 | 0.017 |  |  |  |  |
|  | PBT | 0.260 | 2.752 | 0.007 |  |  |  |  |
|  | z IR | 0.246 | 2.499 | 0.014 |  |  |  |  |
| #3. Insomnia | **Model** | 0.340 | 3.581 | <0.001 | 0.239 | 13.66 | 2/87 | <0.001 |
|  | CRP | 0.295 | 3.102 | 0.003 |  |  |  |  |
|  | z PBT – z SpO2 |  |  |  |  |  |  |  |
| #4. Pure Anxiety | **Model** |  |  |  | 0.365 | 12.20 | 4/85 | <0.001 |
|  | z KYN/TRY | 0.367 | 4.220 | <0.001 |  |  |  |  |
|  | CRP | 0.278 | 3.320 | 0.001 |  |  |  |  |
|  | z duration | 0.314 | 3.583 | <0.001 |  |  |  |  |
|  | BMI | 0.214 | 2.456 | 0.016 |  |  |  |  |
| #5. Pure Depression | **Model** |  |  |  | 0.370 | 25.58 | 2/87 | <0.001 |
|  | CRP | 0.526 | 6.085 | <0.001 |  |  |  |  |
|  | z PBT – z SpO2 | 0.225 | 2.604 | 0.011 |  |  |  |  |
| #6. Physiosomatic symptoms | **Model** |  |  |  | 0.410 | 19.90 | 3/86 | <0.001 |
|  | CRP | 0.349 | 4.16 | <0.001 |  |  |  |  |
|  | z PBT + z duration – z SpO2 | 0.419 | 4.87 | <0.001 |  |  |  |  |
|  | z KYN/TRY | 0.195 | 2.29 | 0.025 |  |  |  |  |
| **#7. Physio-affective**<br>**phenome** | **Model** |  |  |  | 0.428 | 21.44 | 3/86 | <0.001 |
|  | CRP | 0.448 | 5.419 | <0.001 |  |  |  |  |
|  | z PBT + z duration – z SpO2 | 0.305 | 3.601 | <0.001 |  |  |  |  |
|  | z KYN/TRY | 0.259 | 3.095 | 0.003 |  |  |  |  |

CRP: C-reactive protein, FBG: fasting blood glucose, IR: insulin resistance, duration: duration of the acute infectious phase, KYN: kynurenine, TRY: tryptophan, sex: dummy variable with 0=female, 1=male

**Figure 4.**
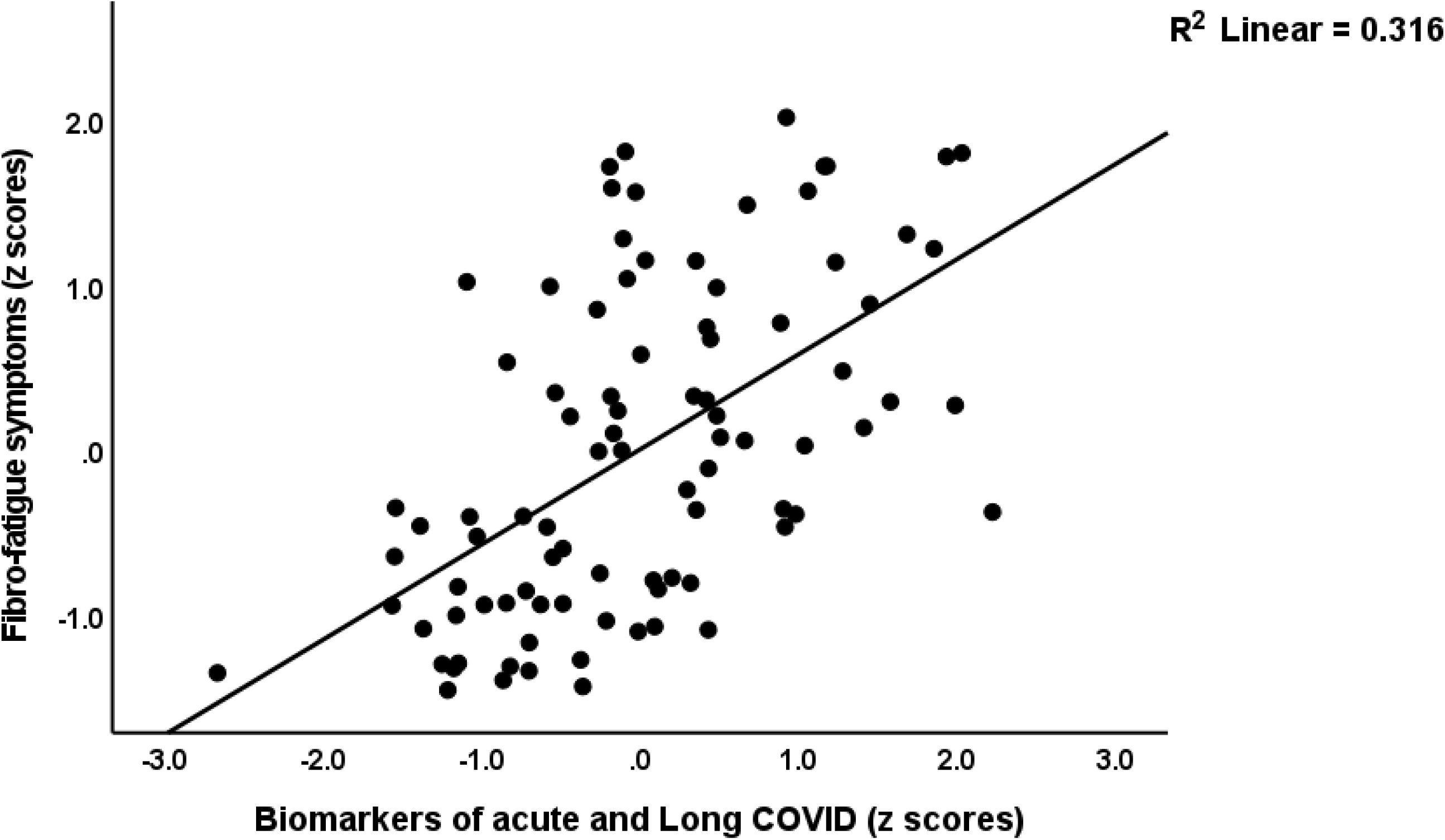
Partial regression of the fibro-fatigue symptoms of Long COVID on the biomarkers of acute COVID-19 (peak body temperature and oxygen saturation) and long COVID (C-reactive protein, the kynurenine/tryptophan ratio, and insulin resistance).

## Discussion

### The physio-affective phenome of Long COVID

The first major finding of this study is that a common core (latent vector) could be extracted from physiosomatic, depressive, and anxiety symptoms, demonstrating that these symptom domains are highly interrelated expressions of the physio-affective phenome of Long COVID. These findings are consistent with our prior findings that a single component drives both physiosomatic and affective symptoms in individuals with Long COVID (Al-Hadrawi, Al-Rubaye et al. 2022, Al-Hakeim, Al-Rubaye et al. 2023, Almulla, Al-Hakeim et al. 2023). In the present investigation, however, the relationships were calculated in patients who had all suffered from acute COVID-19 infection, while in our prior studies, the connections were calculated in healthy controls and participants with long-term COVID. Prior research, including meta-analyses, has generally confirmed that Long COVID is characterized by physiosomatic and affective symptoms (Huang, Huang et al. 2021, Taquet, Geddes et al. 2021, Badenoch, Rengasamy et al. 2022, Premraj, Kannapadi et al. 2022, Titze-de-Almeida, da Cunha et al. 2022). In other illnesses, such as rheumatoid arthritis, major depressive disorder, and schizophrenia, we have previously established intertwined associations between high physiosomatic and affective symptom levels (Kanchanatawan, Sriswasdi et al. 2019, Almulla, Al-Hakeim et al. 2020, Almulla, Al-Rawi et al. 2021, Smesam, Qazmooz et al. 2022).

Previously, we demonstrated that the acute infectious phase of COVID-19 was characterized by increases in physiosomatic and affective symptoms and that a common core could be identified from these domains (Al-Jassas, Al-Hakeim et al. 2022). Hence, both acute COVID-19 and long COVID are linked with significant increases in the physio-affective phenome, indicating that comparable pathways may be responsible for the physio-affective phenome of both acute COVID-19 and long COVID.

We found that 31.1% of COVID-19 patients experienced moderate to severe symptoms of depression consistent with major depression when a cutoff value of 23 on the BDI-II was used (Park, Park et al. 2020). The mean HAMD, HAMA, and FF scores (23.6 ±5.6; 25.4 ±8.6 and 30.9 ±10.1, respectively) for the group of patients with severe COVID indicate that they also suffer from moderate to severe anxiety and mild chronic fatigue syndrome. It is important to note that blood samples were collected between 3 and 12 months (mean: 7.3 ±2.6 months) after remission from the acute phase and that all participants exhibited symptoms that persisted beyond the acute phase of illness, indicating that some Long-term COVID patients developed chronic depressive symptoms, chronic major depression, chronic anxiety disorders, and chronic fatigue syndrome. These results indicate that chronic fatigue and affective symptoms belong to the same symptom spectrum.

In addition, the exclusion of patients with premorbid major depression, anxiety disorders, and chronic fatigue syndrome from our study suggests that some individuals with COVID-19 develop de novo physiosomatic and affective symptoms, as well as major depression and chronic fatigue syndrome, some months later. These findings indicate that both major depression and chronic fatigue syndrome, as well as their comorbidities, may be induced by viral infections. There is now evidence that viral infections are related to the development of chronic fatigue syndrome, severe depression, and anxiety (Maes and Twisk 2010, Bornand, Toovey et al. 2016, Hampton 2016, Karimi, Davoodi et al. 2022).

Similar to earlier studies, the severity of the acute phase inflammatory response (as measured by PBT and SpO2) was substantially linked with the physiosomatic and affective symptoms of acute COVID-19 and long-term COVID, showing that common pathways underlie the acute and long-term COVID phenomes. Specifically, the findings of Al-Hadrawi et al. (2022) and the present results indicate that inflammatory pathways modulate the effects of SARS-CoV-2 infection on the acute and long-term COVID physio-affective phenome (Al-Hadrawi, Al-Rubaye et al. 2022).

### The TRYCAT pathway and Long COVID

The second main conclusion of this research is that severe long-term COVID is related to considerably decreased blood levels of tryptophan and a higher KYN/TRY ratio and that both are associated with anxiety, chronic fatigue, and physiosomatic symptoms. Lower levels of tryptophan and a higher ratio of kynurenine to tryptophan were previously related to somatizing diseases and somatizing symptoms rather than depression per se, and the increased ratio in depression is mostly owing to the presence of somatizing symptoms (Maes and Rief 2012). Hence, one could conclude that the tryptophan-IDO pathway is connected with physiosomatic symptoms rather than depressive symptoms. Yet, as previously mentioned, physiosomatic and affective symptoms are expressions of a shared core (i.e. the phenome), and the KYN/TRY ratio adds to the severity of the physio-affective phenome and is elevated in severe Long COVID.

Our findings that tryptophan is decreased and kynurenine is marginally increased in a cluster of severe Long COVID patients and that the KYN/TRY ratio is associated with the physio-affective phenome of Long COVID (after allowing for the effects of the other biomarkers) suggest that IDO activity may be increased in this patient cluster, albeit with a small effect size. Nonetheless, the correct interpretation of our findings is that tryptophan is probably reduced due to non-IDO-related mechanisms and that, if IDO activity were to increase in any way, IDO self-inhibition has prevented further activation of the TRYCAT pathway in Long COVID despite the mild chronic inflammatory process.

As a substrate, tryptophan (plasma/serum concentration of approximately 12 ng/mL) is readily accessible for kynurenine production (plasma/serum concentration of around 3.3 ng/mL). As a result of the decline in tryptophan in long-term COVID patients, the IDO enzyme may self-regulate and become inactive (Nelp, Kates et al. 2018), and catalytically inactive ferric IDO1 may accumulate and autooxidize (Booth, Basran et al. 2015). In moderately chronic inflammatory circumstances, the latter is likely a self-regulating negative feedback loop that attenuates the creation of neurotoxic TRYCATs. In acute inflammatory and infectious circumstances, tryptophan starvation as a result of cytokine-induced tryptophan depletion is an essential component of the innate immune response that protects against invasive infections and hyperinflammation (Maes, Leonard et al. 2011, Almulla and Maes 2022). Nevertheless, increased kynurenine synthesis produces depressogenic, anxious, excitotoxic, and neurotoxic consequences (Wichers, Koek et al. 2005, Maes 2011). Hence, elevated kynurenine levels may contribute to the physio-affective phenome, which explains why the KYN/TRY ratio is a more accurate index than tryptophan alone.

In recent meta-analyses, no notable changes in TRYCAT production have been observed in depression and schizophrenia, even though low tryptophan levels are a characteristic of both diseases (Almulla, Thipakorn et al. 2022, Almulla, Vasupanrajit et al. 2022). Increased production of TRYCATs appears to be a characteristic of acute inflammatory conditions, such as critical acute COVID-19 infection (Almulla, Al-Hakeim et al. 2023) and interferon-induced depression (Bonaccorso, Marino et al. 2002), as opposed to mild chronic inflammatory conditions, such as depression, schizophrenia, and Alzheimer’s disease (Almulla, Supasitthumrong et al. 2022, Almulla, Thipakorn et al. 2022, Almulla, Vasupanrajit et al. 2022). Reduced blood albumin, which binds tryptophan, is likely the most significant mechanism leading to decreased tryptophan in moderate chronic inflammatory diseases (such as depression and schizophrenia) (McMenamy and Oncley 1958, Yuwiler, Oldendorf et al. 1977, Maes, Minner et al. 1991, Maes, Wauters et al. 1996, Maes, Verkerk et al. 1997). As the availability of plasma tryptophan controls tryptophan levels in the brain (Fernstrom and Wurtman 1972, Partridge 1979), low levels of peripheral tryptophan may contribute to physio-affective symptoms by decreasing central serotonin signaling and decreasing neuroprotective mechanisms (Maes, Galecki et al. 2011, Almulla, Thipakorn et al. 2022). In any event, our data indicate that elevated levels of neurotoxic TRYCATs other than kynurenine do not play a significant role in the establishment of long-term COVID, thereby contradicting findings in severe COVID-19 infection (Almulla, Supasitthumrong et al. 2022).

Our findings further indicate that the intensity of the inflammatory response during acute COVID-19 infection (as measured by a rise in PBT and a decrease in SpO2) is predictive of increases in the KYN/TRY ratio during chronic COVID. Hypoxia may reduce the intracellular concentration of serotonin in lung neuroendocrine cells, and the severity of hypoxia influences the extent of serotonin release (Cutz, Speirs et al. 1993). Consequently, hypoxia during the acute phase of infection may influence the processes of tryptophan depletion and conversion to serotonin.

### Biomarkers of Long COVID

The third major finding of this study is that the three biomarkers of Long COVID assessed in our study (CRP, KYN/TRY, and IR) or acute COVID-19 + Long COVID (CRP, KYN/TRY, IR, SpO2, PBT) together explain a large part of the variance in the physio-affective phenome of Long COVID (around 40-41%). As such, our findings indicate that the inflammatory processes during acute COVID-19 and the combination of immune-inflammatory processes (CRP, KYN/TRP and IR) determine to a large extent the physio-affective phenome of Long COVID. Previously, we reported that immune-inflammatory processes (CRP, activation of the NLRP-3 inflammasome, neuro-oxidative pathways), and IR predict the physio-affective phenome of Long COVID (Al-Hadrawi, Al-Rubaye et al. 2022, Al-Hakeim, Al-Rubaye et al. 2023, Al-Hakeim, Al-Rubaye et al. 2023), and that inflammatory processes predict the physio-affective phenome of acute SARS-CoVv-2 infection (Al-Jassas, Al-Hakeim et al. 2022). Previously, we have discussed the mechanistic processes that may explain the effects of these different pathways on the onset of physio-affective symptoms, with as major culprits increased neurotoxicity of immune products (including kynurenine), neuro-oxidative stress, and increased IR, and lowered neuroprotection by lowered levels of protective antioxidants and tryptophan (Al-Hadrawi, Al-Rubaye et al. 2022, Al-Jassas, Al-Hakeim et al. 2022, Al-Hakeim, Al-Rubaye et al. 2023, Al-Hakeim, Al-Rubaye et al. 2023).

Nevertheless, even more important is our findings that one validated latent construct could be extracted from the three major symptom domains and the composite constructed using the 5 acute and Long COVID biomarkers (CRP, KYN/TRY, IR, SpO2, PBT). This indicates that the physio-affective phenome of Long-COVID is the clinical manifestation of immune-inflammatory processes and new onset IR. Likewise, we were able to construct a new endophenotypic cluster of Long COVID patients with extremely low SpO2, increased kynurenine and very low tryptophan (and thus increased KYN/TRY ratio), very high CRP, moderate increases in IR, and very high ratings on all symptom domains. Lowered SpO2 during the acute phase is therefore a key factor in the onset of severe Long COVID.

## Limitations

This article would have been more interesting if we had measured the cytokines known to affect IDO, such as interferons and IL-1β, as well as reactive oxygen species, which may also increase IDO. One may argue that a sample size of 90 is quite small, decreasing the accuracy of the parameter estimates. Nevertheless, a priori computation of the effect size revealed that a sample size of at least 70 is required to perform multiple regression analyses (the primary outcome of our study) assuming an effect size of 0.2, an alpha of 0.05, and a power of 0.80. In addition, post-hoc analysis performed on the results of our regression studies revealed that even for the regression with the lowest effect size (see table 4, cognitive impairments), we operated with a power of 0.991, and that in all other regressions, we attained a power of 1.

## Conclusions

We were able to develop an endophenotypic class of severe Long COVID (22% of all patients) with very low SpO2 during acute infection, and Long COVID biomarkers including low tryptophan, and elevated kynurenine, KYN/TRY ratio, CRP, and a very high score on physiosomatic, chronic fatigue, depression and anxiety symptom domains. These domains are all indicators of the shared physio-affective phenome of Long COVID. Around 40% of the variation in this phenome was explained by the Long COVID biomarkers, CRP, KYN/TRY, and IR. The physio-affective phenome of Long COVID is a manifestation of inflammatory reactions during acute and Long COVID and decreased plasma tryptophan, whilst increased kynurenine may potentially contribute to these effects.

## Data Availability

The database generated during this study will be made available from the
corresponding author (MM) upon reasonable request once the data set has been fully
exploited by the authors.

## Acknowledgment

We thank the staff of the Dialysis Unit at Al-Hakeem general hospital and Al-Sader medical city in Najaf governorate-Iraq for their help in the collection of samples.

## Funding

This work was supported by an FF66 grant and a Sompoch Endowment Fund from the Faculty of Medicine, MDCU (RA66/016) to MM.

## Conflict of interest

The authors have no conflicts of interest with any industrial or other association concerning the submitted article.

## Author’s contributions

All the contributing authors have participated in the preparation of the manuscript.

## Ethical statement

All controls and patients, or their parents/legal guardians, gave written informed permission before participation in the research. The research was approved by the University of Kufa’s institutional ethics board (8298/2022).

## Data availability statement

The database generated during this study will be made available from the corresponding author (MM) upon reasonable request once the data set has been fully exploited by the authors.

## Notes

### Competing Interest Statement

The authors have declared no competing interest.

